# Structural outcomes among patients receiving targeted therapy for ATTR cardiac amyloidosis - A Systematic Review and Meta-Analysis

**DOI:** 10.64898/2026.08.07.26359992

**Authors:** Revati Varma, Shubhashis Saha, Shreyas Nandyal, Ayodeji Ilelaboye, Saketh Vinjamuri, Aviral Vij, Saurabh Malhotra

**Affiliations:** Department of Cardiology; Department of Internal Medicine, John H. Stroger Hospital, Chicago, IL; Department of Internal Medicine, Gandhi Medical College - Secunderabad, TS, India

**Keywords:** Global longitudinal strain, Interventricular septal thickness, TTR stabilizers, RNA silencers

## Abstract

**Background:** Targeted pharmacologic therapies for transthyretin amyloid cardiomyopathy (ATTR-CM) improve survival; however, their effects on cardiac structural parameters remain incompletely defined.

**Objectives:** To evaluate the pooled effects of disease-modifying therapies for ATTR-CM on echocardiographic structural parameters.

**Methods:** In accordance with PRISMA guidelines, we performed a systematic review and meta-analysis of randomized controlled trials and observational studies published through March 2025 assessing transthyretin stabilizers and RNA-silencing therapies in adults with cardiac amyloidosis. Outcomes included changes in global longitudinal strain (GLS), left ventricular ejection fraction (LVEF), interventricular septal (IVS) thickness, left ventricular mass, stroke volume, E/e′ ratio, and LV end-diastolic volume. Pooled between-group mean differences were calculated using random-effects models. Sensitivity analyses were performed.

**Results:** Eighteen studies (11 randomized, 7 observational) encompassing 3,646 patients were included. Compared with control, drug therapy was associated with attenuation of GLS decline (mean difference [MD] -0.69%; 95% CI -1.10 to -0.29; *P*<0.001) and preservation of LVEF (MD 1.62%; 95% CI 0.73 to 2.51; *P*<0.001). Treatment was also associated with reduced worsening of E/e′ ratio, and preservation of stroke volume. No significant between-group differences were observed for IVS thickness, LV mass and LV end-diastolic volume. Within-group analyses showed no change in echocardiographic parameters between baseline and follow-up in treated patients, in contrast to significant worsening in the control cohort.

**Conclusions:** Disease-modifying therapies for ATTR-CM are associated with stabilization and attenuated progression of cardiac remodeling rather than reversal of structural abnormalities.

## Introduction

Transthyretin amyloid cardiomyopathy (ATTR-CM) is an infiltrative cardiomyopathy, where abnormal amyloid fibrils accumulate within heart muscle tissue. The progressive accumulation of amyloid fibrils compromises cardiac function, leading to heart failure and increased mortality. Recently, there have been significant advances in targeted ATTR-CM pharmacologic therapies aimed at slowing disease progression. Transthyretin silencers and stabilizers are now approved and available for treatment of patients with ATTR-CM, and have been shown to improve survival (Refs – ATTR-ACT, ATTRIBUTE-CM and HELIOS-B).

Disease-modifying therapies for ATTR-CM act upstream of amyloid fibril formation – either via messenger RNA silencing mechanisms in the liver or by stabilization of circulating TTR tetramer. Despite a direct mechanistic link between these drugs’ mechanism of action and their ability to influence cardiac function, there has been growing interest in measuring secondary benefits on cardiac structure and function from these novel therapies. However, studies to date have been inconsistent in their findings, have used inconsistent methodologies for assessing cardiac structure or function, or have focused on a single treatment type. This meta-analysis was performed to assess the pooled effects of targeted drug therapies for ATTR-CM on structural echocardiographic parameters in patients with ATTR-CM.

## Methods

In accordance with the Preferred Reporting Items for Systematic Reviews and Meta-Analyses (PRISMA) guidelines,^1^ we conducted a systematic review and meta-analysis to assess the use of drug therapy (TTR stabilizers like tafamidis, and acoramidis, small interfering RNAs like patisiran, vutrisiran and revusiran, antisense oligonucleotides like inotersen and eplontersen) on outcomes in patients with CA. No data involving human participants were collected, and therefore institutional ethical/IRB approval was not required. All included studies report obtaining their own ethical approval and/or participant consent as stated in their original publications. The review protocol was registered with the International Prospective Register of Systematic Reviews (PROSPERO; registration number CRD420251183198). The registered protocol prespecified three outcome domains: survival and cardiovascular hospitalization, functional capacity and quality of life, and echocardiographic measures of cardiac structure and function. The present report addresses the echocardiographic domain.

### Study search strategy and literature search

A systematic search of literature of studies published from inception to March 2025 was conducted using PubMed, Cochrane and Ovid MEDLINE databases. We included both randomized controlled trials (RCTs) as well as observational studies in the analysis. The search strategy included terms such as “Cardiac amyloidosis” OR “CA” OR “Amyloidosis” OR “transthyretin” or “ATTR” or “ATTR-CM” OR “patisiran” OR “vutrisiran” OR “tafamidis” OR “Acoramidis” OR “eplontersen” OR “Inotersen” OR “RNA silencing.” Both medical Subject Headings (MeSH) and title/abstract keywords were included in the search strategy. There were no restrictions on the language or geographical region where the study was conducted.

All the above-mentioned databases were searched independently using the search strategy, and resulting studies were combined and entered into the Rayyan software. Duplicates were removed. In the initial step, two investigators (SN and RV) independently reviewed headings and abstracts to either include or exclude studies based on the selection criteria. In the second round, full-text manuscripts were reviewed independently, and initially selected studies were further included or excluded based on the agreed-upon criteria. This was followed by a third round of resolving conflicts among the selected studies. This systematic and thorough approach advanced the robustness of our search and helped minimize potential biases. After the final selection and inclusion of papers, the data were collected and synthesized by one investigator and then cross-checked by the second investigator for accuracy.

### Inclusion and exclusion criteria

Studies (including both RCTs and observational studies) were included in the meta-analysis if they met the following criteria: (1) adult population > 18 years of age with an established diagnosis of CA. For studies conducted on patients with systemic amyloidosis, we included data from the cardiac subpopulation only; (2) intervention included any of the drug therapies mentioned above; (3) must have a comparator group which could be placebo or treatment of choice (universally named as the control group); (4) studies required at least one of the following outcomes - echocardiographic endpoints such as ejection fraction (EF), global longitudinal strain (GLS), Left ventricular mass (LV Mass), interventricular septal thickness (IVS thickness), LV End diastolic volume (LVEDV), or stroke volume. Exclusion criteria included (1) patients <18 years of age, (2) studies that did not consider CA or did not have a cardiac subpopulation. For studies evaluating multiple dosing regimens, the FDA-approved dose was selected when available; otherwise, the higher dose was used for analysis. For non-FDA-approved therapies, the higher dose was chosen based on the assumption that greater dosing may be associated with improved outcomes. Two investigators independently reviewed the abstracts from the initial search to exclude studies that did not meet the above criteria and then conducted a second round of scrutinizing full-text articles. All conflicts were resolved with a third investigator (SV).

### Data extraction and outcomes

We extracted general information for each study. This included the title of the study, first author, year of publication, study design, sample size, intervention and control group, clinical characteristics of the participants, outcome data assessed in each study, and the duration of follow up. Outcomes included echocardiographic endpoints such as ejection fraction (EF), global longitudinal strain (GLS), left ventricular mass (LV Mass), interventricular septal thickness (IVS thickness), LV End diastolic volume (LVEDV), and stroke volume. The change between baseline and follow-up in continuous outcomes was reported as means and standard deviations (SD). When reported, least square means and standard errors, medians and interquartile ranges or medians and ranges, were converted into mean (SD) using established conversion methods.^2^

### Risk of bias and quality assessment

For RCTs, the RoB-2 tool was used to assess the risk of bias. Cochrane domains were used in assessing RoB. Each study was assigned a RoB score in 6 Cochrane domains. The six domains included ‘Random sequence generation,’ ‘Allocation concealment,’ ‘Blinding of the participants and personnel,’ ‘Blinding of outcome assessment,’ ‘Incomplete outcome data,’ ‘Selective reporting.’ The RoB scales included low risk, moderate or unclear risk and high risk. Low risk was assigned a score of 1, moderate risk a score of 2, and high risk a score of 3. The highest score in each domain was considered as the overall RoB score for that study.

For non-randomized observational studies, risk of bias was assessed using the ROBINS-I (Risk Of Bias In Non-randomized Studies of Interventions) tool.^3^ Bias was evaluated across seven domains. These include bias due to confounding, selection of participants, classification of interventions, missing data, measurement of outcomes, and selection of the reported result. Each domain was graded as low, moderate, serious or critical risk of bias. The overall risk of bias for each study was determined by the highest level of risk identified in any of the seven domains.

### Statistical analysis

All data were entered into an Excel sheet and statistical analysis was performed using RStudio. For continuous outcomes, pooled effect estimates were calculated using the between-group mean difference (MD) and 95% confidence intervals (CI). A random effects model was applied to account for between-study heterogeneity. The restricted maximum-likelihood (REML) method was used to calculate the between-study variance. CIs were estimated using the DerSimonian–Laird framework. Heterogeneity was quantified using the I² statistic. The robustness of the pooled estimates was assessed using a leave-one-out sensitivity analysis, in which each study was sequentially excluded and the analysis repeated to assess whether any single study disproportionately influenced the overall effect size.

Forest plots displayed the mean change (follow-up-baseline) in the intervention and control groups separately, as well as the between-group difference (mean change in the intervention group - mean change in the control group). Individual between-group difference for each study and the pooled mean difference across all studies and 95% CI was depicted for each outcome. A p-value <0.05 was considered significant. A prespecified subgroup analysis was performed by stratifying studies by design (RCTs and observational studies) to assess consistency between real-world and trial-based evidence. These analyses were performed using stratified random effects models. Separate forest plots were generated for each subgroup. Additionally, the change from baseline to follow-up within each cohort was depicted using histograms. For this, we included only studies that reported baseline and follow-up data as mean (SD). Within-group changes were compared using a paired t-test.

## Results

### Baseline demographics

This meta-analysis included a total of 18 studies, including RCTs and observational studies, evaluating drugs for CA. Eleven studies were RCTs and 7 were observational studies. Nine studies evaluated tafamidis, 3 evaluated patisiran, 2 evaluated vutrisiran, and one each evaluated inotersen, eplontersen, revusiran and coramitug (Table 1). Rettl, et al. studied two doses of tafamidis (61 mg and 20 mg), of which the data from 60 mg tafamidis was used for analysis. Fontana et al. studied two doses of coramitug (10 and 60 mg), of which data from 60 mg coramitug was used. Of the 11 RCTs, 7 were judged to have a low RoB, 3 moderate and 1 with a high RoB. Of the 7 observational studies, 3 had moderate and 4 had serious risk of bias (Figure 2). Across all studies, the total number of patients treated with drug therapy was 2080, and 1566 were either untreated with the study drug or were given placebo. The mean age among patients receiving the intervention and control was respectively 71± 8 years and 73± 8 years.

**Table 1.** Characteristics of included randomized and observational studies assessing treatment options for cardiac amyloidosis.

| Study | Year | Intervention | Type of study | Dose | Control | Intervention, n | Control, n |
| --- | --- | --- | --- | --- | --- | --- | --- |
| Benson, et al.<sup>13</sup> | 2018 | Inotersen | RCT | 300 mg | Placebo | 112 | 60 |
| Chamling, et al.<sup>14</sup> | 2022 | Tafamidis | Retrospective observational trial | 61 mg | Untreated | 20 | 20 |
| Dobner, et al.<sup>15</sup> | 2024 | Tafamidis | Prospective observational study | 61 mg | Untreated | 36 | 15 |
| Fontana, et al.<sup>16</sup> | 2024 | Vutrisiran | RCT | 25 mg | Placebo | 326 | 328 |
| Giblin, et al.<sup>17</sup> | 2022 | Tafamidis | Retrospective observational trial | 61 mg | Untreated | 23 | 22 |
| Judge Daniel, et al.<sup>18</sup> | 2020 | Revusiran | RCT | 500 mg | Placebo | 140 | 60 |
| Kuyama, et al.<sup>19</sup> | 2025 | Tafamidis | Retrospective observational trial | 20 mg | Untreated | 151 | 107 |
| Masri, et al.<sup>20</sup> | 2024 | Eplontersen | RCT | 45 mg | Placebo | 49 | 30 |
| Maurer, et al.<sup>21</sup> | 2018 | Tafamidis | RCT | Pooled | Placebo | 181 | 179 |
| Restelli, et al.<sup>23</sup> | 2025 | Tafamidis | Retrospective observational trial | 61 mg | Untreated | 28 | 11 |
| Rettl, et al.<sup>24</sup> | 2022 | Tafamidis | Observational study | 61 mg | Untreated | 35 | 19 |
| Rosenblum, et al.<sup>25</sup> | 2023 | Patisiran | RCT | 0.3 mg/kg | Placebo | 90 | 36 |
| Shah, et al.<sup>4</sup> | 2024 | Tafamidis | RCT | 80 mg | Placebo | 176 | 177 |
| Solomon, et al.<sup>27</sup> | 2019 | Patisiran | RCT | 0.3 mg/kg | Placebo | 90 | 36 |
| Takashio, et al.<sup>28</sup> | 2023 | Tafamidis | Retrospective observational trial | 80 mg | Untreated | 125 | 55 |
| Jering, et al.<sup>29</sup> | 2025 | Vutrisiran | RCT | 25 mg | Placebo | 196 | 199 |
| Maurer et al.<sup>30</sup> | 2023 | Patisiran | RCT | 0.3 mg/kg | Placebo | 267 | 177 |
| Fontana, et al.-<br>Coramitug 60<sup>31</sup> | 2025 | Coramitug | RCT | 60 mg/Kg | Placebo | 35 | 35 |

Abbreviations. RCT, randomized controlled trial

**Figure 1.**
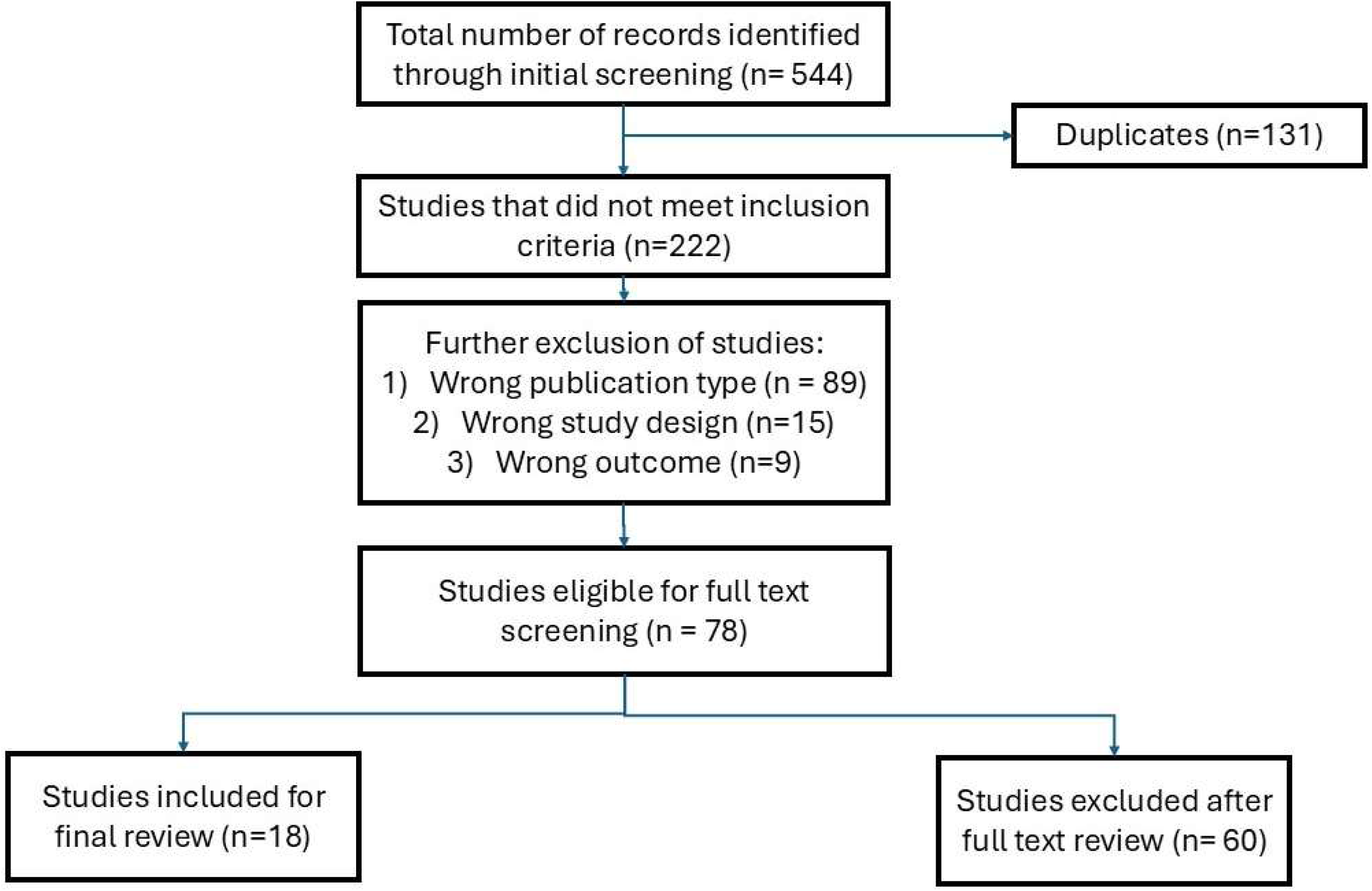
PRISMA diagram depicting selection and screening of studies. Out of 544 studies selected after initial screening, 78 were eligible for full text review. Of these. 60 were excluded, leaving a total of 18 studies included for final review.

**Figure 2.**
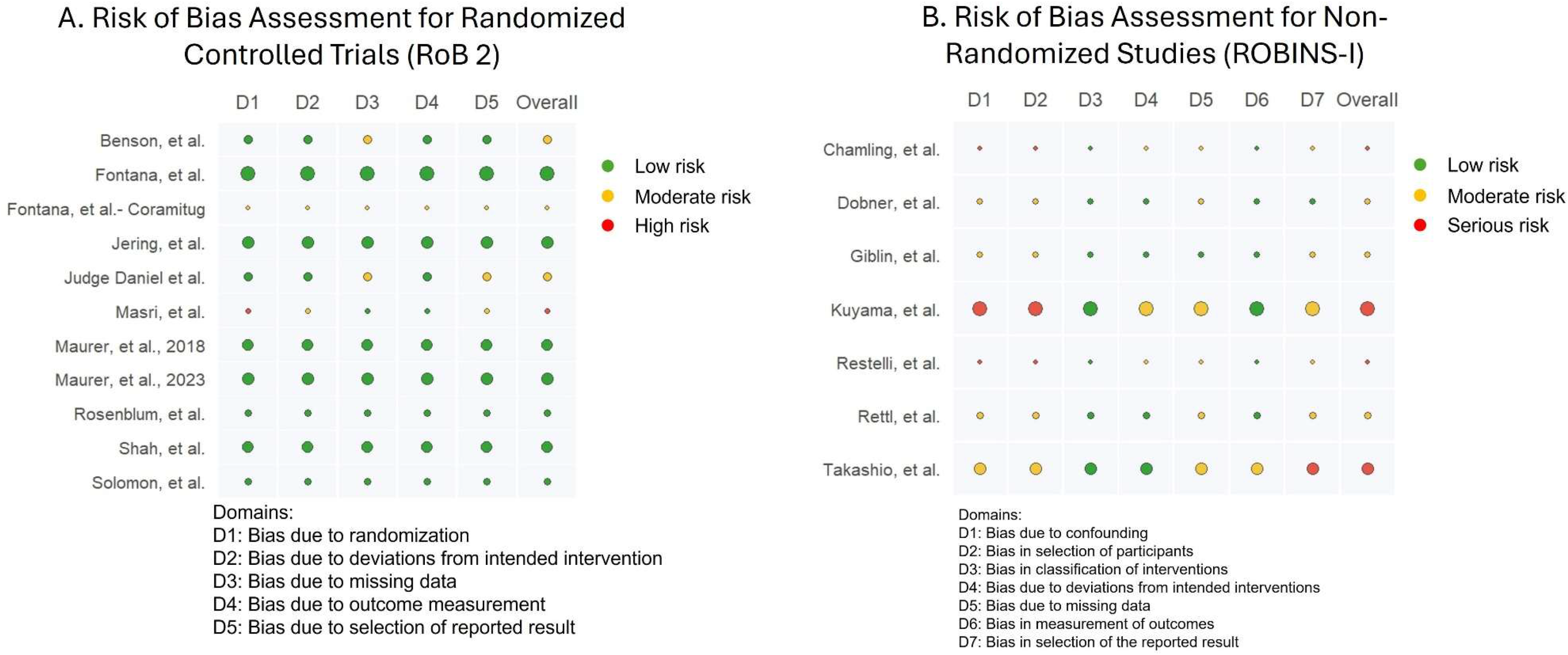
Summary of Risk of Bias Assessments for Randomized and Non-Randomized Studies. For figure A, the green, yellow and red circles represent low, intermediate and high risk of bias. For figure B, the corresponding colors represent low, moderate and serious risk of bias. The size of the circles represents the sample size of the study.

### Global Longitudinal Strain

There were a total of 14 studies that evaluated GLS (n = 1626 in the intervention group and n= 1332 in the control group). The average duration of follow-up was 22 months. In the random effects model, drug therapy attenuated the decline in GLS compared to control (MD= - 0.69% CI -1.10 to -0.29; P<0.001). Subgroup analysis based on the type of study showed that the initial significant difference disappeared when only RCTs were included (MD= -0.45% CI -1.11 to -0.21; P=0.183), while it remained with observational studies alone: MD= -0.92% CI -1.28 to -0.56; P<0.001). Findings were consistent even after sequentially omitting studies in the sensitivity analysis. Five studies reported baseline and follow up values as mean (SD). There was no difference in the GLS at baseline compared to follow up in the intervention group (−10.18 (2.64) vs -9.89 (2.48); P= 0.267), while there was a significant worsening of GLS in the control group (−10.64 (1.58) vs -9.41 (1.49); P= 0.003) (Figure 3).

**Figure 3.**
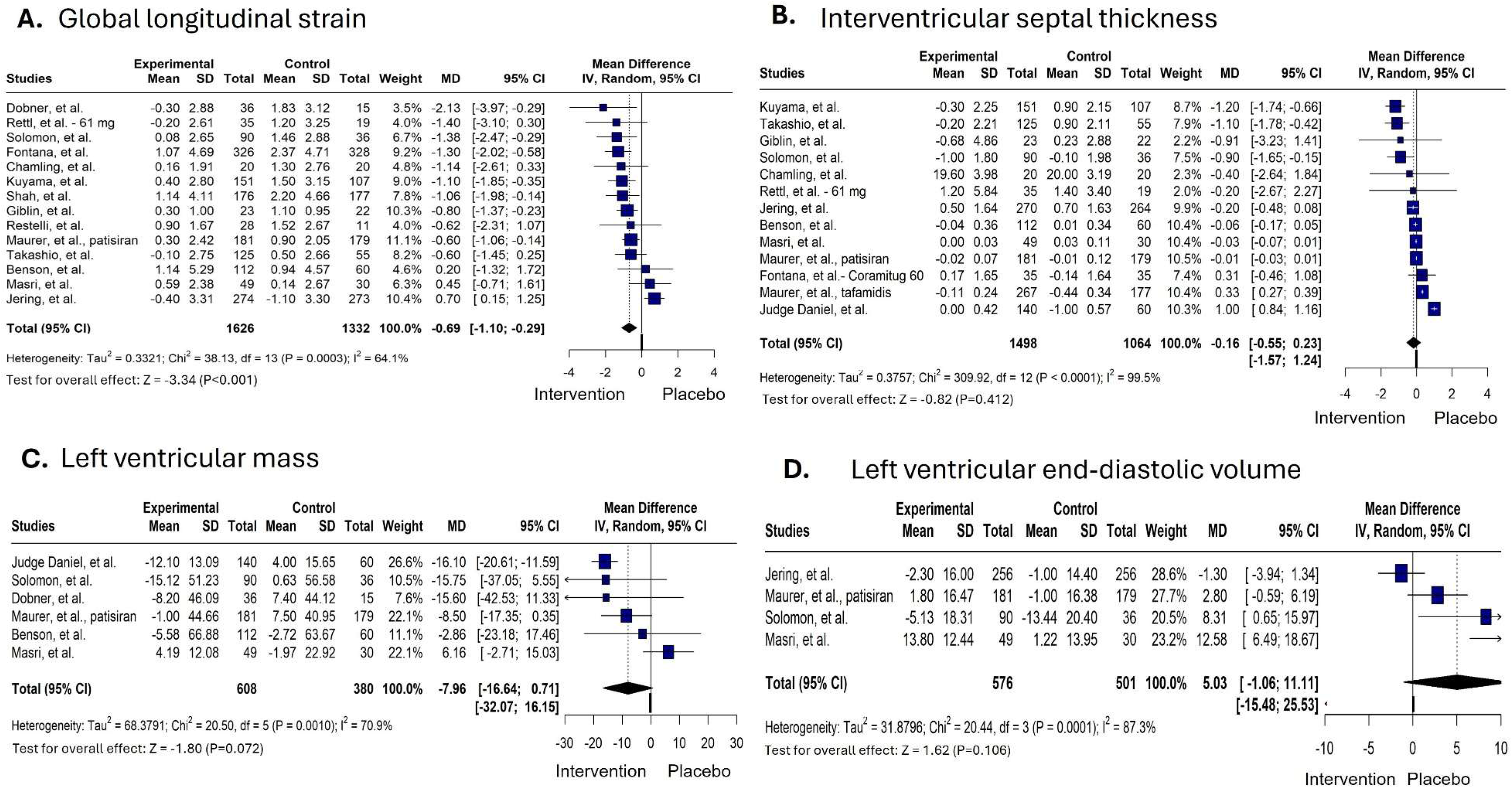
Forest plot depicting change in GLS, IVS, LV mass and LVEDV. Between-group mean difference for each study is reported. The pooled between-group mean difference is depicted with the 95% CI.

### Interventricular septal thickness

There were 13 studies that evaluated IVS thickness (n= 1498 in the intervention group and n= 1064 in the control group). The mean duration of follow-up was 21 months. There was no difference in the magnitude of decline in the IVS thickness between treatment and placebo groups (MD -0.16 mm; 95% CI, -0.55 to 0.23; P=0.412). Subgroup analysis using only RCTs was consistent with the overall findings (MD 0.09 mm; 95% CI, -0.25 to 0.44; P=0.590), but using observational studies alone, there was a significant difference ( MD -1.10 mm; 95% CI, - 1.51 to -0.70; P <001). The sensitivity analysis did not reveal a single study contributing to observed results. Among seven studies, there was no difference in the IVS thickness at baseline compared to follow up both in the intervention group (20.77 (9.23) vs 20.93 (9.61); P= 0.563), as well as in the control group (16.88 (2.41) vs 17.31 (2.50); P= 0.207) (Figure 3).

### Left Ventricular Ejection Fraction

A total of 12 studies evaluated the drug therapy on ejection fraction (n=1189 in the intervention group, n=832 in the control group). Mean duration of follow up was 20 months. Drug therapy was associated with preservation of EF (MD=1.62 percentage points; 95% CI 0.73 to 2.51; P <0.001). When analyzing separately by RCTs or by observational studies, preservation of LVEF was noted in the treatment group in both cases. Sensitivity analysis did not yield a single study driving the results. While there was a significant decline in the EF from baseline to follow up in the control group (53.34 (2.93) % vs 49.63 (2.25) %; P= 0.007), there was no change in the intervention group (51.30 (4.20) vs 50.07 (2.92); P= 0.283) (Figure 4).

**Figure 4.**
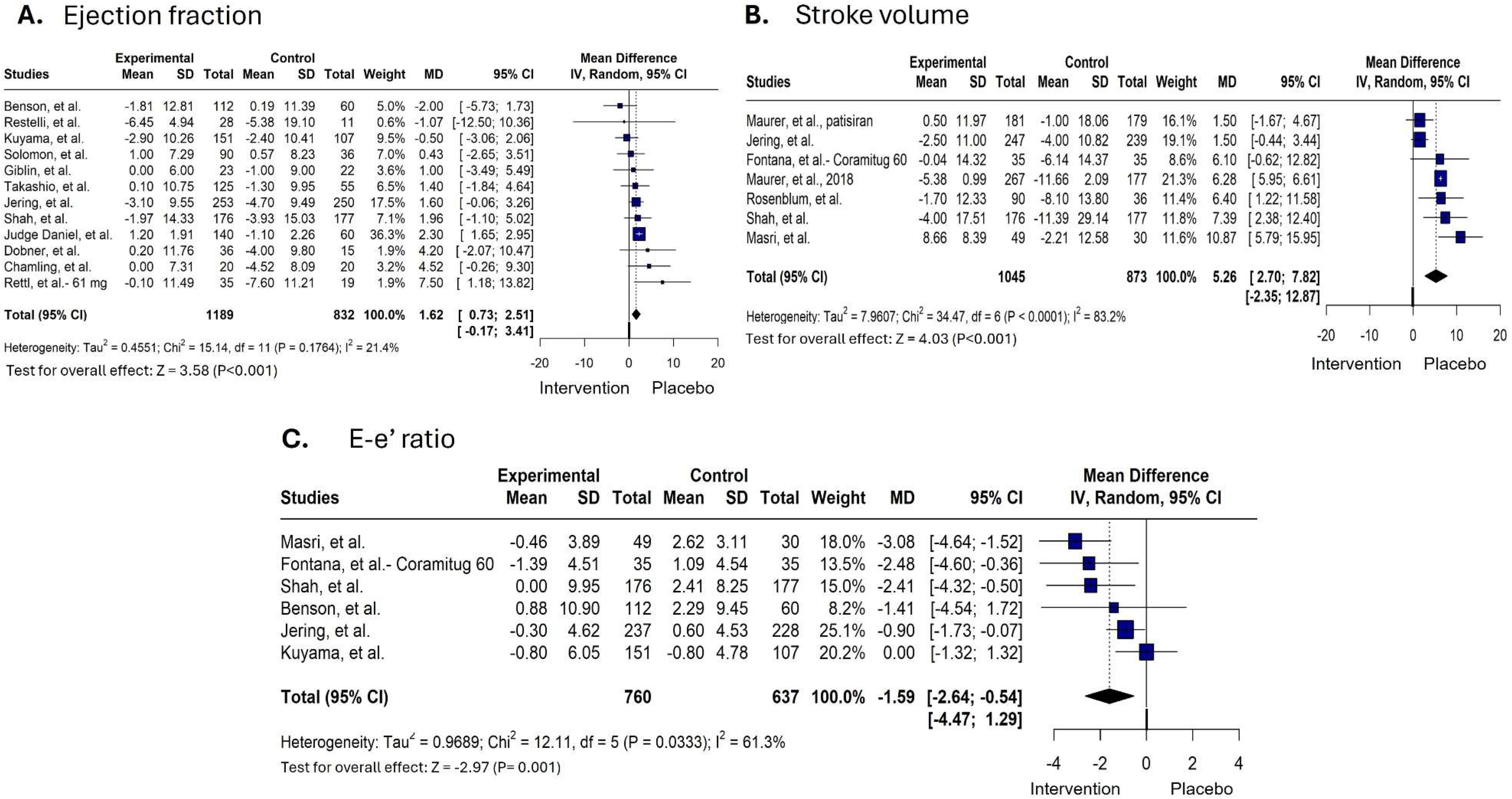
Forest plot depicting change in ejection fraction, stroke volume and E/e’ ratio. Forest plot depicting change in ejection fraction (A), stroke volume (B) and E/e’ ratio (C). Between-group mean difference for each study is mentioned. The pooled between-group mean difference is depicted with the 95% CI.

**Figure 5.**
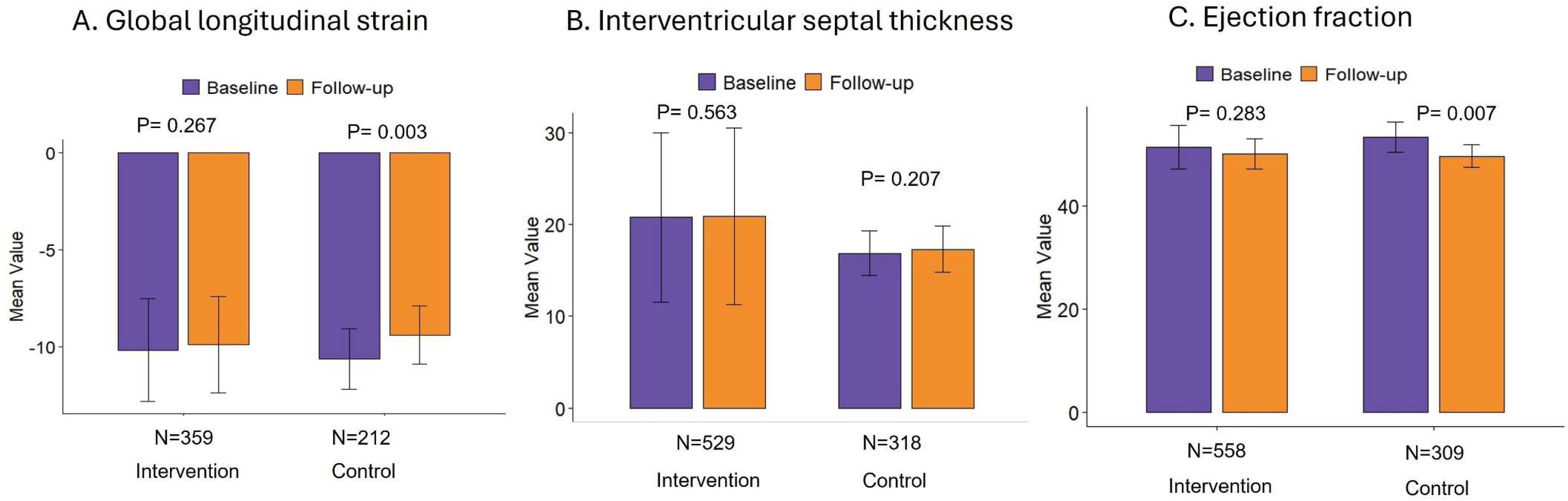
Histogram of follow up and baseline mean (SD) of cardiac structural parameters. Notice that cardiac structural parameters remained unchanged in the intervention group but worsened significantly in the control group.

### Other end points

The treatment group was associated with preservation of E/e’ ratio (MD -1.59; 95% CI, - 2.64 to -0.54; P= 0.001), and stroke volume (MD 5.26 mL,95% CI, 2.70 to 7.82; P <0.001), when compared to the control group. However, there was no effect of treatment on LV mass (MD -7.96 grams; 95% CI, -16.64 to 0.71; P =0.072), and LVEDV (MD 5.03 mL; 95% CI, -1.06 to 11.11; P =0.106).

## Discussion

In this meta-analysis of 18 studies evaluating disease-modifying therapies for transthyretin cardiac amyloidosis, treatment with TTR stabilizers and silencers was associated with preservation of several echocardiographic markers of cardiac performance, including LVEF, GLS, stroke volume, and E/e’. In contrast, no significant changes were observed in LV wall thickness, LV mass, or LV end-diastolic volume. Taken together, these findings suggest that currently available ATTR-CM therapies may slow functional deterioration and hemodynamic progression even when overt reverse structural remodeling is not apparent within currently studied follow-up periods.

Over the last several years, the treatment landscape for ATTR-CM has evolved rapidly with the development of therapies targeting amyloidogenesis upstream of TTR fibril formation. Tafamidis and acoramidis act by stabilizing the circulating TTR tetramer^4,5^, whereas RNA-targeted therapies reduce hepatic production of mutant and wild-type TTR^6,7^. Although major randomized trials have demonstrated improvements in mortality, hospitalization, and functional capacity,^8,9^ less is known regarding the extent to which these therapies alter cardiac structure and function. Prior studies examining echocardiographic endpoints have yielded inconsistent findings, likely due to differences in patient populations, imaging protocols, therapeutic classes, and follow-up durations. The current analysis was therefore designed to evaluate the collective effect of ATTR-directed therapies on cardiac structure and function, as measured by echocardiography, across both clinical trials and real-world observational cohorts.

One notable finding was the relative preservation of GLS in treated patients. Longitudinal strain is particularly relevant in cardiac amyloidosis because impairment in longitudinal mechanics frequently precedes changes in conventional systolic function.^10^ Progressive worsening in GLS likely reflects advancing myocardial infiltration and worsening myocardial deformation. In the present analysis, patients treated with targeted ATTR therapies demonstrated attenuated decline in GLS when compared with controls; however, this effect was not significant when analyses were restricted to randomized trials alone. Several factors may explain this observation. Variability in strain acquisition and vendor-specific analysis platforms remains an important limitation in pooled echocardiographic studies.^11^ In addition, the degree of change in GLS over short follow-up intervals may be modest and difficult to detect in controlled trial settings. Nevertheless, the within-group analyses are clinically informative, demonstrating relative stability in GLS among treated patients while untreated cohorts experienced progressive worsening over time.

A similar pattern was observed with LVEF. Although LVEF is often preserved until later stages of ATTR-CM, deterioration in EF generally reflects more advanced cardiomyopathy and carries adverse prognostic implications.^10^ In our analysis, targeted therapy was associated with maintenance of LVEF, whereas untreated patients demonstrated a significant decline during follow-up. The magnitude of improvement was modest but is biologically consistent with the expected mechanism of current therapies. Existing ATTR-directed therapies primarily reduce ongoing amyloid fibril formation and deposition rather than directly remove established myocardial fibrils.^4,6^ Accordingly, stabilization rather than normalization of ventricular function may represent the more realistic therapeutic effect over currently available follow-up durations.

The observed preservation in stroke volume and E/e’ ratio further supports a favorable physiologic effect of therapy. ATTR-CM is characterized by restrictive ventricular physiology with progressive impairment in filling dynamics and declining forward cardiac output.^10^ Maintenance of stroke volume may therefore reflect stabilization of ventricular performance and may partially explain improvements in exercise tolerance and clinical status reported in major therapeutic trials.^8,9^ Similarly, attenuation of worsening filling pressures suggests a potential slowing of progressive ventricular stiffening and diastolic dysfunction. These findings emphasize that meaningful therapeutic benefit may occur even in the absence of measurable reductions in myocardial wall thickness.

Importantly, no significant treatment effect was observed for IVS thickness, LV mass, or LVEDV. While TTR silencers and stabilizers reduce the formation and deposition of transthyretin amyloid fibrils, they are not designed to clear pre-existing amyloid fibrils from the myocardium.^4,6^ Structural echocardiographic parameters may therefore remain relatively unchanged despite stabilization in myocardial performance. In addition, myocardial thickening in ATTR-CM reflects not only amyloid infiltration but also extracellular matrix expansion and chronic remodeling,^12,13^ processes that may not reverse rapidly. It is possible that substantially longer follow-up durations or future fibril-clearing therapies will be required before consistent regression in myocardial wall thickness or mass can be demonstrated.^14^

The discordance between functional stabilization and limited structural change may have implications for surveillance imaging in ATTR-CM. Functional markers such as GLS, stroke volume, and filling pressures may be more sensitive indicators of therapeutic response than conventional structural measurements alone. These observations also support the growing role of multimodality imaging approaches incorporating echocardiography, cardiac magnetic resonance imaging, and nuclear scintigraphy to better characterize disease progression and treatment response.^15^

Several limitations should be acknowledged. Included studies were heterogeneous with respect to design, therapeutic agent, imaging methodology, and follow-up duration. A number of pooled findings were influenced by observational studies, which remain susceptible to residual confounding and selection bias. Echocardiographic assessment, particularly GLS, may vary across software platforms and with variability in image acquisition. In addition, follow-up durations may not have been long enough to capture meaningful structural remodeling. Therapies with differing mechanisms of action were analyzed together, limiting the ability to determine whether certain drug classes exert differential effects on myocardial structure or function. Finally, echocardiographic parameters may not be adequate to ascertain changes in amyloid burden, which could be measured by advanced cardiac magnetic resonance and radionuclide imaging.

In conclusion, the present analysis provides a comprehensive assessment of echocardiographic changes associated with modern ATTR-directed therapies. The findings suggest that current treatments primarily act by slowing the progression of myocardial dysfunction and preserving cardiac performance rather than reversing established structural disease. As newer combination strategies and fibril-clearing therapies continue to emerge, future studies with longer follow-up and standardized imaging protocols will be important in determining whether true reverse remodeling can ultimately be achieved in ATTR-CM.

## Clinical Perspectives

In patients with ATTR-CM, disease-modifying therapies are associated with stabilization of cardiac structure and an attenuated decline in certain functional parameters, with no significant reversal of myocardial remodeling. These findings highlight that current therapies primarily improve outcomes by preserving cardiac function. Stabilization of cardiac function over time could be used as a marker of therapeutic benefit.

## Data Availability

All data extracted for this meta-analysis are derived from previously published studies, which are cited within the manuscript. The full data extraction spreadsheet used in the pooled analysis is available from the corresponding author upon reasonable request.

## Central Illustration

**Figure 1.**
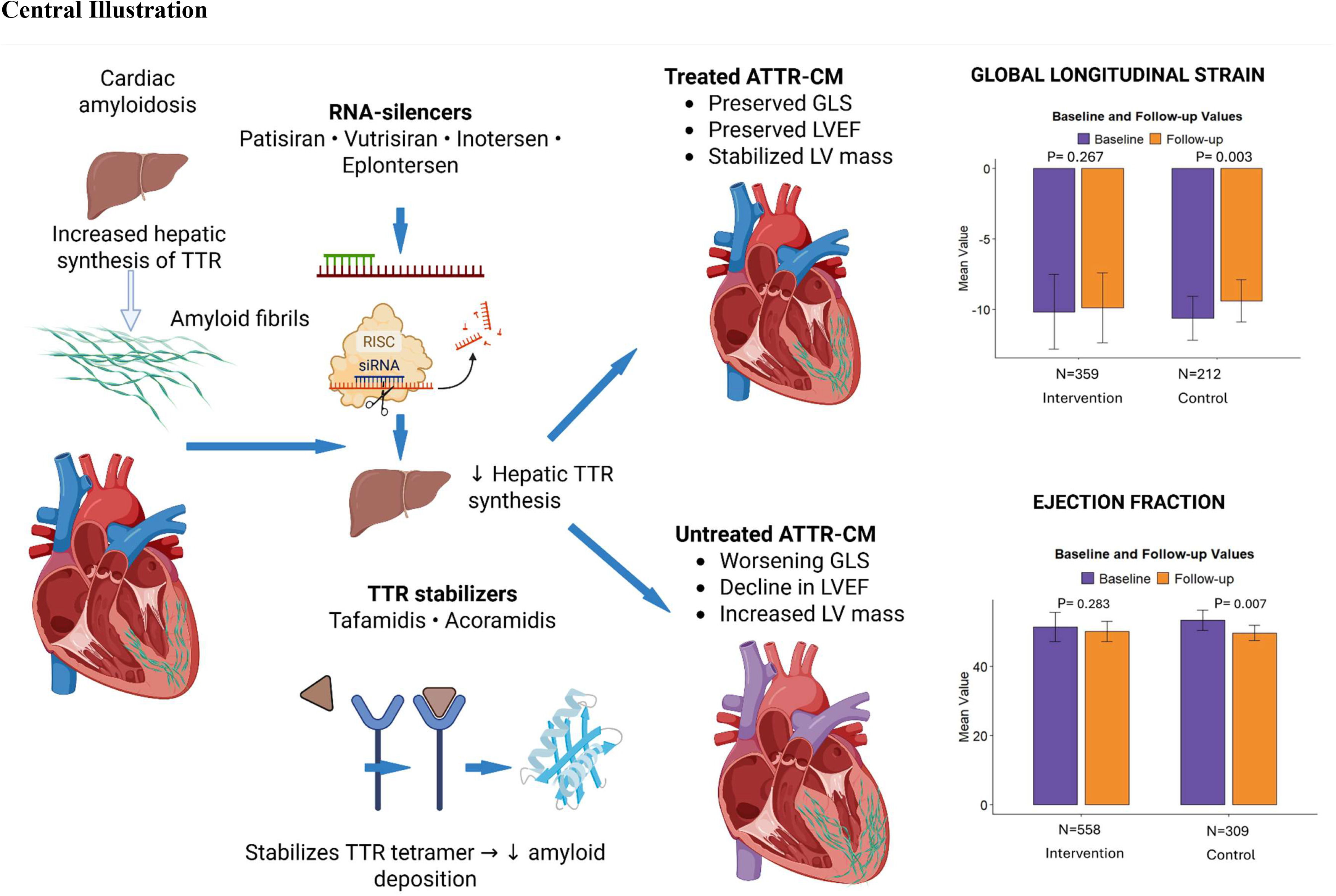
Central Illustration. Disease-modifying therapies for ATTR-CM, including RNA-silencing agents and transthyretin stabilizers, act upstream to reduce amyloid progression. While these therapies do not reverse established myocardial remodeling, they are associated with stabilization of cardiac structural parameters, in contrast to progressive deterioration observed in untreated patients.

## References

1. Page MJ, Moher D, Bossuyt PM, et al. PRISMA 2020 explanation and elaboration: updated guidance and exemplars for reporting systematic reviews. BMJ. 2021;372:n160. doi:10.1136/bmj.n160

2. Wan X, Wang W, Liu J, Tong T. Estimating the sample mean and standard deviation from the sample size, median, range and/or interquartile range. BMC Med Res Methodol. 2014;14:135. doi:10.1186/1471-2288-14-135

3. Sterne JA, Hernán MA, Reeves BC, et al. ROBINS-I: a tool for assessing risk of bias in non-randomised studies of interventions. BMJ. 2016;355:i4919. doi:10.1136/bmj.i4919

4. Bulawa CE, Connelly S, Devit M, et al. Tafamidis, a potent and selective transthyretin kinetic stabilizer that inhibits the amyloid cascade. Proc Natl Acad Sci U S A. 2012;109(24):9629–9634. doi:10.1073/pnas.1121005109

5. Gillmore JD, Judge DP, Cappelli F, et al. Efficacy and Safety of Acoramidis in Transthyretin Amyloid Cardiomyopathy. N Engl J Med. 2024;390(2):132–142. doi:10.1056/NEJMoa2305434

6. Adams D, Gonzalez-Duarte A, O’Riordan WD, et al. Patisiran, an RNAi Therapeutic, for Hereditary Transthyretin Amyloidosis. N Engl J Med. 2018;379(1):11–21. doi:10.1056/NEJMoa1716153

7. Benson MD, Waddington-Cruz M, Berk JL, et al. Inotersen Treatment for Patients with Hereditary Transthyretin Amyloidosis. N Engl J Med. 2018;379(1):22–31. doi:10.1056/NEJMoa1716793

8. Maurer MS, Schwartz JH, Gundapaneni B, et al. Tafamidis Treatment for Patients with Transthyretin Amyloid Cardiomyopathy. N Engl J Med. 2018;379(11):1007–1016. doi:10.1056/NEJMoa1805689

9. Fontana M, Berk JL, Gillmore JD, et al. Vutrisiran in Patients with Transthyretin Amyloidosis with Cardiomyopathy. N Engl J Med. 2025;392(1):33–44. doi:10.1056/NEJMoa2409134

10. Kittleson MM, Maurer MS, Ambardekar AV, et al. Cardiac Amyloidosis: Evolving Diagnosis and Management: A Scientific Statement From the American Heart Association. Circulation. 2020;142:e7–e22. doi:10.1161/CIR.0000000000000792

11. Farsalinos KE, Daraban AM, Ünlü S, Thomas JD, Badano LP, Voigt JU. Head-to-Head Comparison of Global Longitudinal Strain Measurements among Nine Different Vendors: The EACVI/ASE Inter-Vendor Comparison Study. J Am Soc Echocardiogr. 2015;28(10):1171–1181.e2. doi:10.1016/j.echo.2015.06.011

12. Chamling B, Bietenbeck M, Korthals D, et al. Therapeutic value of tafamidis in patients with wild-type transthyretin amyloidosis (ATTRwt) with cardiomyopathy based on cardiovascular magnetic resonance (CMR) imaging. Clin Res Cardiol. 2023;112(3):353–362. doi:10.1007/s00392-022-02035-w

13. Dobner S, Bernhard B, Ninck L, et al. Impact of tafamidis on myocardial function and CMR tissue characteristics in transthyretin amyloid cardiomyopathy. ESC Heart Fail. 2024;11(5):2759–2768. doi:10.1002/ehf2.14815

14. Fontana M, Garcia-Pavia P, Grogan M, et al. Coramitug, a Humanized Monoclonal Antibody for the Treatment of Transthyretin Amyloid Cardiomyopathy: a Phase 2, Randomized, Multicenter, Double-Blind, Placebo-Controlled Trial. Circulation. Published online November 10, 2025. doi:10.1161/CIRCULATIONAHA.125.077304

15. Dorbala S, Ando Y, Bokhari S, et al. ASNC/AHA/ASE/EANM/HFSA/ISA/SCMR/SNMMI Expert Consensus Recommendations for Multimodality Imaging in Cardiac Amyloidosis: Part 1 of 2—Evidence Base and Standardized Methods of Imaging. Circ Cardiovasc Imaging. 2021;14:e000029. doi:10.1161/HCI.0000000000000029

